# InterMEL: An international biorepository and clinical database to uncover predictors of survival in early-stage melanoma

**DOI:** 10.1101/2022.05.21.22275329

**Authors:** Irene Orlow, Keimya D. Sadeghi, Sharon N. Edmiston, Jessica M. Kenney, Cecilia Lezcano, James S. Wilmott, Anne E. Cust, Richard A. Scolyer, Graham J. Mann, Tim K. Lee, Hazel Burke, Valerie Jakrot, Ping Shang, Peter M. Ferguson, Tawny W. Boyce, Jennifer S. Ko, Peter Ngo, Pauline Funchain, Judy R. Rees, Kelli O’Connell, Honglin Hao, Eloise Parrish, Kathleen Conway, Paul B. Googe, David W. Ollila, Stergios J. Moschos, Eva Hernando, Douglas Hanniford, Diana Argibay, Christopher I. Amos, Jeffrey E. Lee, Iman Osman, Li Luo, Pei-Fen Kuan, Arshi Aurora, Bonnie E. Gould Rothberg, Marcus W. Bosenberg, Meg R. Gerstenblith, Cheryl Thompson, Paul N. Bogner, Ivan P. Gorlov, Sheri L. Holmen, Elise K. Brunsgaard, Yvonne M. Saenger, Ronglai Shen, Venkatraman Seshan, Eduardo Nagore, Marc S. Ernstoff, Klaus J. Busam, Colin B. Begg, Nancy E. Thomas, Marianne Berwick, the InterMEL Consortium

**Author notes:** Corresponding author (IO). Membership of the InterMEL Consortium is provided in the Acknowledgements.

## Abstract

**Introduction:** We are conducting a multicenter study to identify classifiers predictive of disease-specific survival in patients with primary melanomas. Here we delineate the unique aspects, challenges, and best practices for optimizing a study of generally small-sized pigmented tumor samples including primary melanomas of at least 1.05mm from AJTCC TNM stage IIA-IIID patients. This ongoing study will target 1,000 melanomas within the international *InterMEL* consortium. We also evaluated tissue-derived predictors of extracted nucleic acids’ quality and success in downstream testing.

**Methods:** Following a pre-established protocol, participating centers ship formalin-fixed paraffin embedded (FFPE) tissue sections to Memorial Sloan Kettering Cancer Center for the centralized handling, dermatopathology review and histology-guided coextraction of RNA and DNA. Samples are distributed for evaluation of somatic mutations using next gen sequencing (NGS) with the MSK-IMPACT™ assay, methylation-profiling (array), and miRNA expression (Nanostring nCounter).

**Results:** Sufficient material was obtained for screening of miRNA expression in 683/685 (99%) eligible melanomas, methylation in 467 (68%), and somatic mutations in 560 (82%). In 446/685 (65%) cases, aliquots of RNA/DNA were sufficient for testing with all three platforms. Among samples evaluated by the time of this analysis, the mean NGS coverage was 249x, 59 (18.6%) samples had coverage below 100x, and 41/414 (10%) failed methylation QC due to low intensity probes or insufficient Meta-Mixed Interquartile (BMIQ)- and single sample (ss)- Noob normalizations. Six of 683 RNAs (1%) failed Nanostring QC due to the low proportion of probes above the minimum threshold. Age of the FFPE tissue blocks (p<0.001) and time elapsed from sectioning to co-extraction (p=0.002) were associated with methylation screening failures. Melanin reduced the ability to amplify fragments of 200bp or greater (absent/lightly pigmented vs heavily pigmented, p<0.003). Conversely, heavily pigmented tumors rendered greater amounts of RNA (p<0.001), and of RNA above 200 nucleotides (p<0.001).

**Conclusion:** Our experience with many archival tissues demonstrates that with careful management of tissue processing and quality control it is possible to conduct multi-omic studies in a complex multi-institutional setting for investigations involving minute quantities of FFPE tumors, as in studies of early-stage melanoma.

## Introduction

Melanoma accounts for the great majority of deaths from skin cancer with an estimated 7,650 deaths in the USA in 2022 (1). The five-year survival rate of melanoma ranges from 98% for localized disease to less than 30% in patients with distant metastases at the time of diagnosis (2, 3). Factors known to affect progression and survival include primary tumor characteristics (4, 5), presence of nodal or distant metastases at the time of diagnosis (6), anatomic site of the tumor (7), as well as demographic characteristics such as age at diagnosis (8, 9) and sex (9); however, the prediction of individual outcomes based on clinicopathologic factors has limited power. The addition of molecular characteristics has the potential to improve risk classification and help identify individuals more likely to benefit from a differential surveillance schedule and/or therapies.

To date, the most comprehensive efforts undertaken to reveal the multifaceted molecular characteristics of cutaneous melanoma were reported by The Cancer Genome Atlas (TCGA) (10). These investigations led to the classification of melanomas into four subtypes based on somatic *BRAF, RAS, and NF1* mutations, and the identification of phenotypic characteristics (i.e., tumor infiltrating lymphocytes and LCK protein expression) linked to improved survival. The number of primary melanomas in TCGA, however, was small (n∼67) and the thickness of these tumors was larger than the vast majority of primary melanomas, which are usually very thin at diagnosis. Thus, findings may not be generalizable. To be able to translate research findings into the real clinical and diagnostic practice, it is necessary to utilize melanoma tissue that remains available once histopathologic diagnosis is completed as part of the standard care. Therefore, for findings to be generalizable, investigations need to focus on the use of limited archival tissue from small melanomas.

In this article we describe our experiences organizing the tissue collection and processing of specimens that are being collected by the multi-institutional InterMEL Consortium (11), a study designed to define molecular subtypes of cutaneous primary melanoma of at least 1.05 mm in patients diagnosed with AJCC TNM stages IIA through IIID melanoma to improve the prognostic stratification, to guide enhanced surveillance, and/or the use of adjuvant therapies for early intervention, as necessary.

The use of targeted and immuno-based therapies for stage III melanoma patients has been shown to be effective in improving recurrence-free survival (12). Also, pembrolizumab has just received FDA approval as adjuvant treatment for stage IIB/C melanoma based on its significant efficacy compared to placebo in preventing both locoregional and distant recurrence in a double-blind clinical trial (13). Therefore, it is pertinent and timely to better identify which patients are at highest risk who would most likely benefit from these treatments and which patients are at lower risk who should be spared of unnecessary, potentially toxic, and costly treatments (14).

The InterMEL Consortium is collecting clinical and pathologic data, as well as archived melanoma tissues, to screen for somatic mutations and evaluate miRNA and mRNA expression, methylation, protein expression, and to then correlate these features with survival. A uniquely challenging aspect of the current study is the use of archived primary melanomas, which are typically of limited size. To address this concern our study design involves the systematic co-extraction of DNA and RNA from the same tissue sections and derived cell lysates. This study design allows us to save tissue, and most importantly, it allows us analysis of molecular data from the same portion of tumor, eliminating potential biases due to intratumor heterogeneity.

In this report we detail procedures involved in establishing the appropriate infrastructure for our multicenter, integrated study. These include tissues and workflow, sample quality and quantity parameters, and criteria for DNA and RNA distribution for testing, for procuring a balanced proportion of controls (survivors for >5 years with no evidence of disease) and cases (dead within 5-years of follow up) across tests. We also report on the various quality control (QC) parameters used. Based on our experience on already procured data and FFPE tissues from 685 eligible early-stage melanomas from 10 centers, we provide insights into the logistic and technical challenges involved when conducting a large patho-epidemiologic international and multicenter study of disease-specific survival in 1,000 primary melanomas from stage II/ III patients.

## Methods

### Overview

This retrospective ongoing study aims to collect biospecimens and data from a total of 1,000 eligible melanoma patients, including patients who died of melanoma within 5 years of diagnosis (referred to as cases) and patients who lived/are alive for at least 5 years after diagnosis, without evidence of regional or distant melanoma recurrence or relapse (referred to as controls). Participants are identified through the multi-center and international consortium ‘Integration of Clinical and Molecular Biomarkers for Melanoma Survival’ or InterMEL (11), as further detailed below. The study protocol was approved by the Institutional Review Boards at each participating institution, material and data user agreements are in place, and research has been conducted according to the principles expressed in the Declaration of Helsinki. The Biospecimen Core provides complete pathology and biospecimens support, and appropriate infrastructure to all investigations. Activities are detailed next and in the Supporting Information.

### Participants

For each participant, the following data elements are being collected by the participating study centers: demographics, vital status, year of diagnosis, follow up time, progression, recurrence, anthropomorphic measures, treatment type. For the index primary tumors, single *versus* multiple status, TNM stage (AJCC Ed. 8), tumor burden or load, and anatomic site are also recorded. The following study centers provided patients’ data and biospecimens as of March 7^th^ 2021: Cleveland Clinic (Cleveland, OH), Case Western Reserve University (Cleveland, OH), Dartmouth Cancer Center (Lebanon, NH), the University of Texas MD Anderson Cancer Center (Houston, TX), Melanoma Institute Australia (Sydney, NSW, Australia), Memorial Sloan Kettering Cancer Center (New York, NY), New York University Langone Medical Center (New York, NY), The University of North Carolina (Chapel Hill, NC, hereafter denoted UNC), Roswell Park Comprehensive Cancer Center (Buffalo, NY),Yale School of Medicine (New Haven, CT). Additional centers are currently preparing data and tissue for participants not included in this report.

### Biospecimens

The study collects archived tumor and counterpart non-tumor tissue (or germline DNA), following a detailed protocol and shipping manifest template for the assembly, labeling, transport and shipping of samples, and communication procedures that are provided in advance to each participating center, with further guidance provided as needed by the Biospecimens and the Administrative Cores. All tumor samples are procured from FFPE tissue blocks sectioned at each of the contributing centers, for this study.

The overall flow of specimens and related information is depicted in Fig 1. First, study centers provide the minimal electronic data required to verify eligibility of cases and controls centrally (T.K.L.), and then eligibility is communicated to the centers, and to the Biospecimens and Administrative Cores. For eligible melanomas, members of the Molecular Epidemiology Laboratory at Memorial Sloan Kettering (MSK) in New York print and mail 2D alphanumeric temperature and solvent resistant labels to the centers. The study centers, in turn, prepare tumor and non-tumor samples following a common Biospecimen Core Tissue Handling and Sectioning Standard Operating Procedure. Detailed procedures and recommendations are provided in the Supporting Information, Appendix 1.

**Fig 1.**
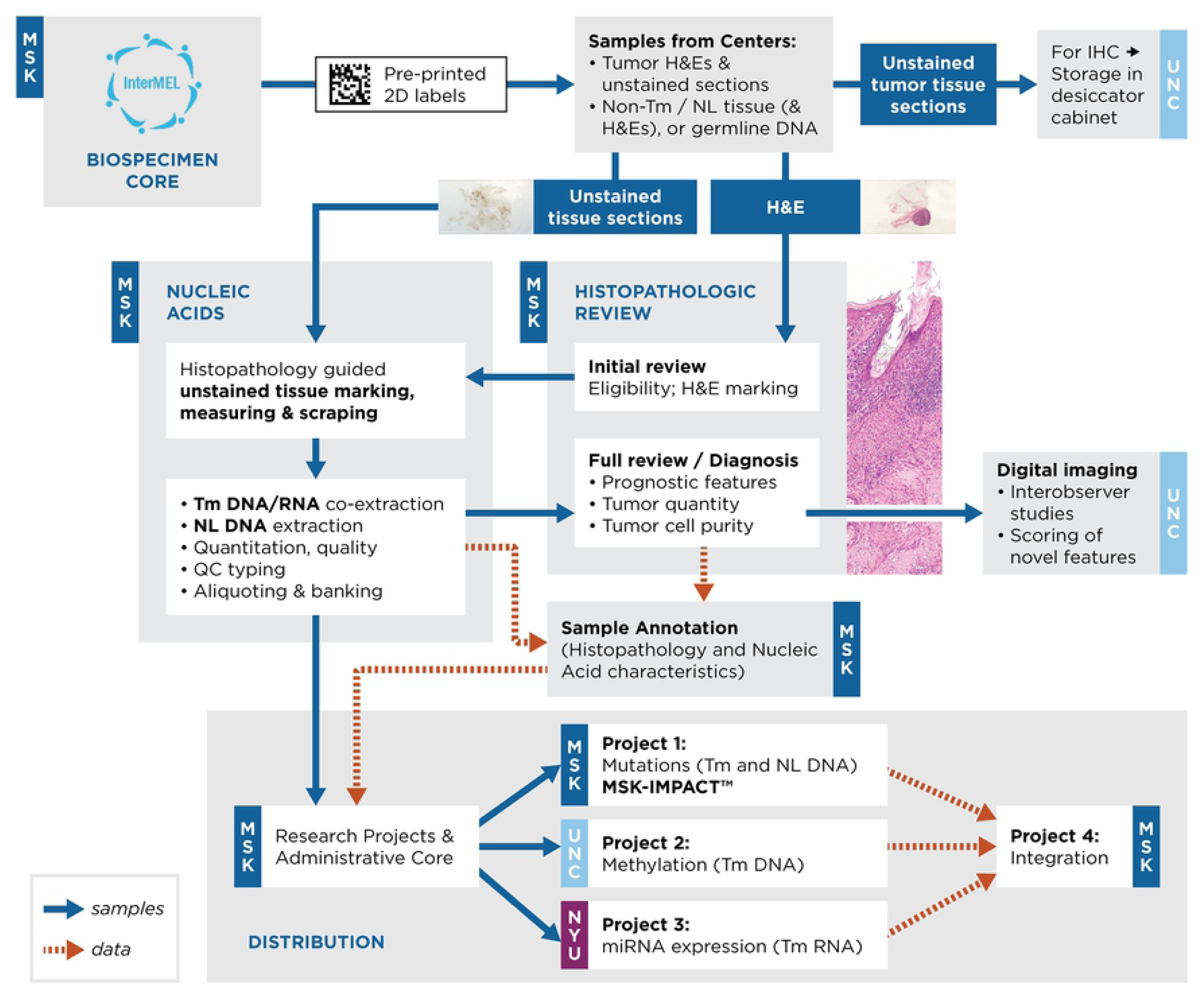
Summary of specimens and data flow in InterMEL.

### Pathology review

Tumor H&Es undergo two rounds of centralized pathology review by expert dermatopathologists at MSK (K.B., C.L.). The initial pathology review serves to confirm the eligibility of the case and the eligibility of the tissue for nucleic acid extractions. The eligible area is marked with a water-resistant pen directly on the H&E. The eligible and marked H&Es are then utilized in the Laboratory as guides for (a) determining and annotating the size of the qualifying tissue area, and (b) for marking the unstained tissue sections in preparation of nucleic acid extraction (see below). Next, the tumor H&Es are returned to the reference pathologist for a second –more in-depth, or ‘full’-pathology review. Here, melanoma diagnosis, histopathologic prognostic features, volume, viability, and purity of available tumor tissue are documented. The annotated histopathologic prognostic features included, at minimum: Breslow thickness, ulceration, melanoma subtype (ICD-0-3 morphology code), Clark level, mitotic rate, tumor infiltrating lymphocytes (TIL) grade, regression, pigmentation, associated nevus, solar elastosis, predominant cell type, neurotropism. Additional salient observations (e.g., whether tissues appeared to be decalcified, necrotic, or include scarring) are also noted. Participants and tumor variables and their definitions are listed in the Supporting Material (Appendix, Supporting Table S1).

### Histopathology-guided co-extraction of nucleic acids from archival tissues

For each tumor specimen, systematic marking of qualifying unstained tissue areas is performed. This is followed by careful scraping of tissues and batched co-extraction of nucleic acids using the AllPrep® DNA/RNA FFPE Kit (Qiagen). Details on the preliminary optimization, co-extraction procedures, and aliquoting rules, can be found in the Supporting Information (S1 Appendix, S1 Fig, S2 Appendix). The extraction kits and procedures utilized for procurement of germline DNA (gDNA), and the detailed procedures utilized for the assessment of DNA and RNA quantity and quality, are also found in the Supporting Information (S1 Appendix). Upon completion of the co-extraction, H&Es are scanned (for future image analysis).

### Criteria for allocation of specimens for testing

Primarily, we use the amount of double stranded (dsDNA) tumor and normal DNA, and total RNA as criteria for distribution for testing. To date, RNA samples with yields of 500ng or greater were distributed for testing. The distribution of DNA involves consideration of quality, quantity and availability of tumor and normal DNA for Next Generation Sequencing (NGS), and a balanced distribution of samples/cases for methylation and NGS. Specifically, we triage study material using the following rules:

a. When equal or greater than (≥) 350ng of tumor DNA and ≥100ng of normal DNA are obtained, DNAs are distributed for <u>both</u> mutation (tumor-normal paired DNAs) and methylation testing (tumor DNA).
b. When 250 to 349ng of tumor DNA and ≥ 100ng of normal DNA are available, DNA samples are distributed for <u>either</u> mutation or methylation testing, procuring a balanced distribution of controls (survivors) and cases (dead within 5-years of follow up) across tests.
c. Samples with less than 70ng of DNA were held back.
d. Participants that lack or have insufficient amounts of germline or non-tumor DNA available are held back until sent as a tumor DNA-only batch for NGS.

When pairing tumor and normal DNAs for NGS, we take into consideration both amounts and quality of the DNA. For example, relatively greater amounts of FFPE tumor DNA are paired with germline DNA extracted from blood (or lower amounts of blood-DNA is paired with tumor DNA). Qualifying specimens are plated and distributed with accompanying documentation for testing of mutations at MSK, methylation at UNC and miRNA profiling at the NYU.

### Screening of somatic mutations, methylation, and miRNA expression

The methodology is described in detail in the Supporting Information, within the S1 Appendix. Briefly, DNA samples are sequenced at Memorial Sloan Kettering using the Integrated <u>M</u>utation <u>P</u>rofiling of <u>A</u>ctionable <u>C</u>ancer <u>T</u>argets or MSK-IMPACT™, a clinically validated and FDA approved hybridization capture-based next-generation sequencing assay developed to guide cancer treatment (15, 16). At the University of North Carolina, DNA samples undergo a bisulfite modification, and genome-wide DNA methylation analysis is performed using the Infinium MethylationEPIC BeadChip kit (Illumina). Filtering and normalization of output methylation data obtained from the Infinium MethylationEPIC arrays is done in R (17). RNA samples are evaluated for miRNA expression at New York University using the Nanostring nCounter® Human v3 miRNA Expression Assay, which includes 800 microRNAs.

### Data handling

The centers providing melanoma specimens prepare an Excel file containing the de-identified information necessary to assess eligibility. This file is uploaded onto a secure cloud where information is checked semi-automatically assisted by a custom-built SAS program. As of March 7th, 2021, 793 Excel records (325 cases and 468 controls) were assessed, and 296 (92%) cases and 413 (88%) controls were deemed eligible for the study. Additional information regarding communication of eligibility, annotation of data, and data sharing, is provided in the Supporting Information (S1 Appendix).

### Statistical analysis

Relationships between continuous variables (e.g., tissue area and DNA yield) were tested using Pearson correlation tests. Fisher’s Exact test was used to evaluate the associations among categorical variables (e.g., survival group and ulceration), and Welch t-test and ANOVA were used to evaluate potential associations between continuous and categorical variables. Tumor purity expressed by the proportion of tumor cells, was dichotomized into >0.7 or ≤0.7 (>70 and ≤70% tumor cells). DNA amplifiability was dichotomized two ways, according to the DNA amplification of two or more fragments, and three or more fragments. Pigmentation (absent, lightly pigmented, or heavily pigmented) and the IGO core DNA and Library QC recommendations (fail, try, pass) were analyzed as three categories, or collapsed into two categories: pigmentation absent/lightly pigmented vs heavily pigmented, and recommendations Fail vs Try/Pass. Associations were illustrated using scatterplots, boxplots, and bar graphs. Statistical analyses were performed in R (17), v.3.6.3.

## Results

### Study participants and melanomas

This report includes the data and analysis for 685/718 eligible melanomas received at MSK as of March 7^th^, 2021. Upon completion of the initial central pathology review, tissue ineligibility was attributed to:

- Inability to confirm a primary melanoma diagnosis on tissue sections provided for centralized review: lymph node metastasis with no definitive primary melanoma; absence of visible skin; presence of mostly congenital, residual, or deep penetrating nevus
- Inadequate tissue quality (e.g., decalcified, mostly scar, or mostly necrotic); and/or quantity (e.g., melanoma size and/or amounts of remaining tumor

Normal tissue was excluded from further analysis when inadequate in size, contaminated with tumor, or when the counterpart tumor was deemed ineligible. Subsequently, 9 confirmed melanoma patients were also excluded because they were lost to follow up and/or had died from unknown causes.

A second centralized and full pathology review was completed in 561/685 melanomas (as of the cut-off date for the current analysis and manuscript), and data are summarized in Table 1. Of these 327 (58.3%) are “controls” (survived >5 years without a melanoma recurrence) and 234 (41.7%) are “cases” (died within the 5-year follow up period). Most participants were reported being male (61.5%), white (95.2%) and had a median age at diagnosis of 64 years. Tumor thickness ranged from 1.1 to 120mm. Two-hundred and eighty-three of the participants (52.2%) presented at diagnosis with stage II and 268 (47.8%) with stage III at diagnosis. Additional variables of interest were captured (Table 1).

**Table 1.**
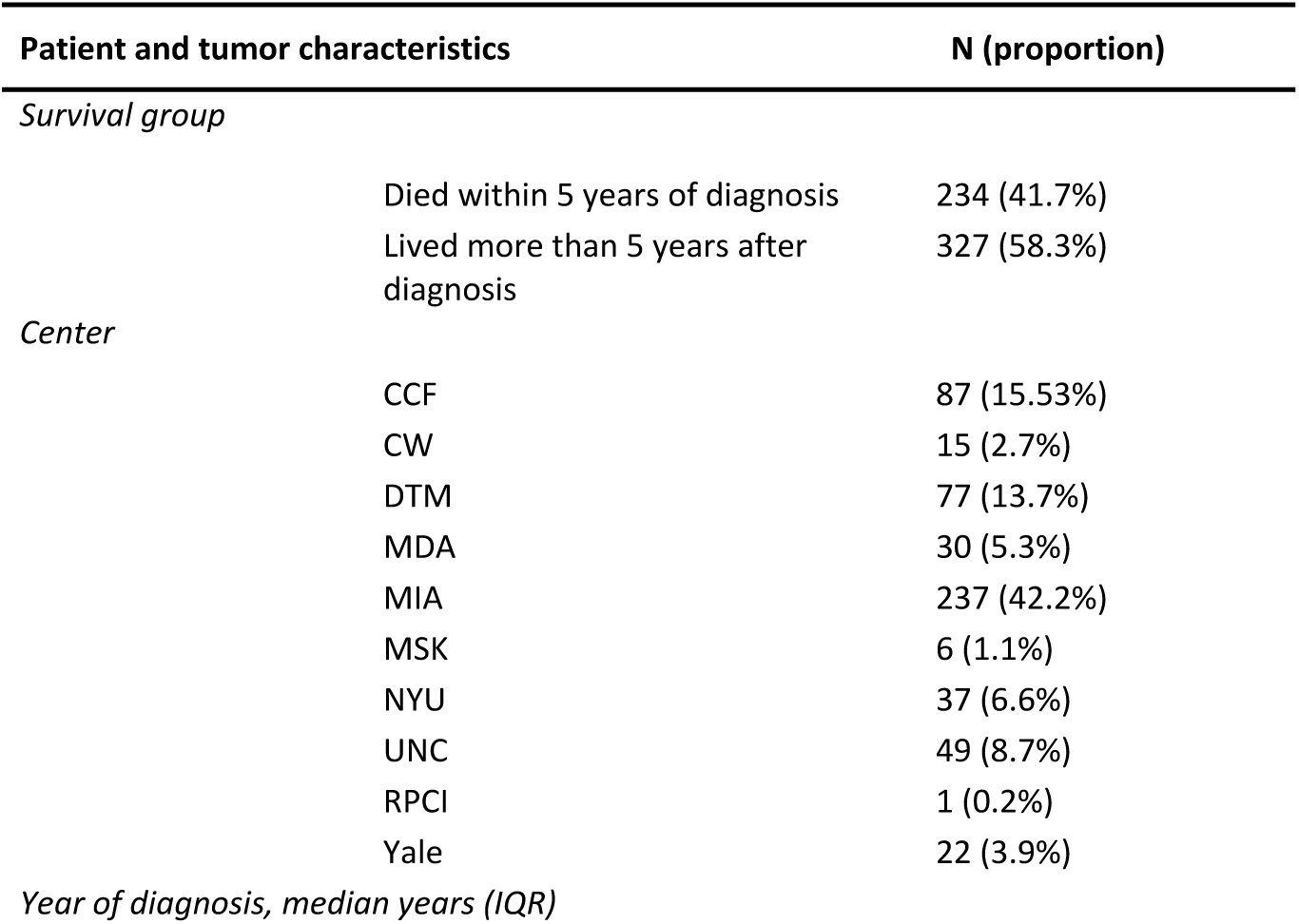
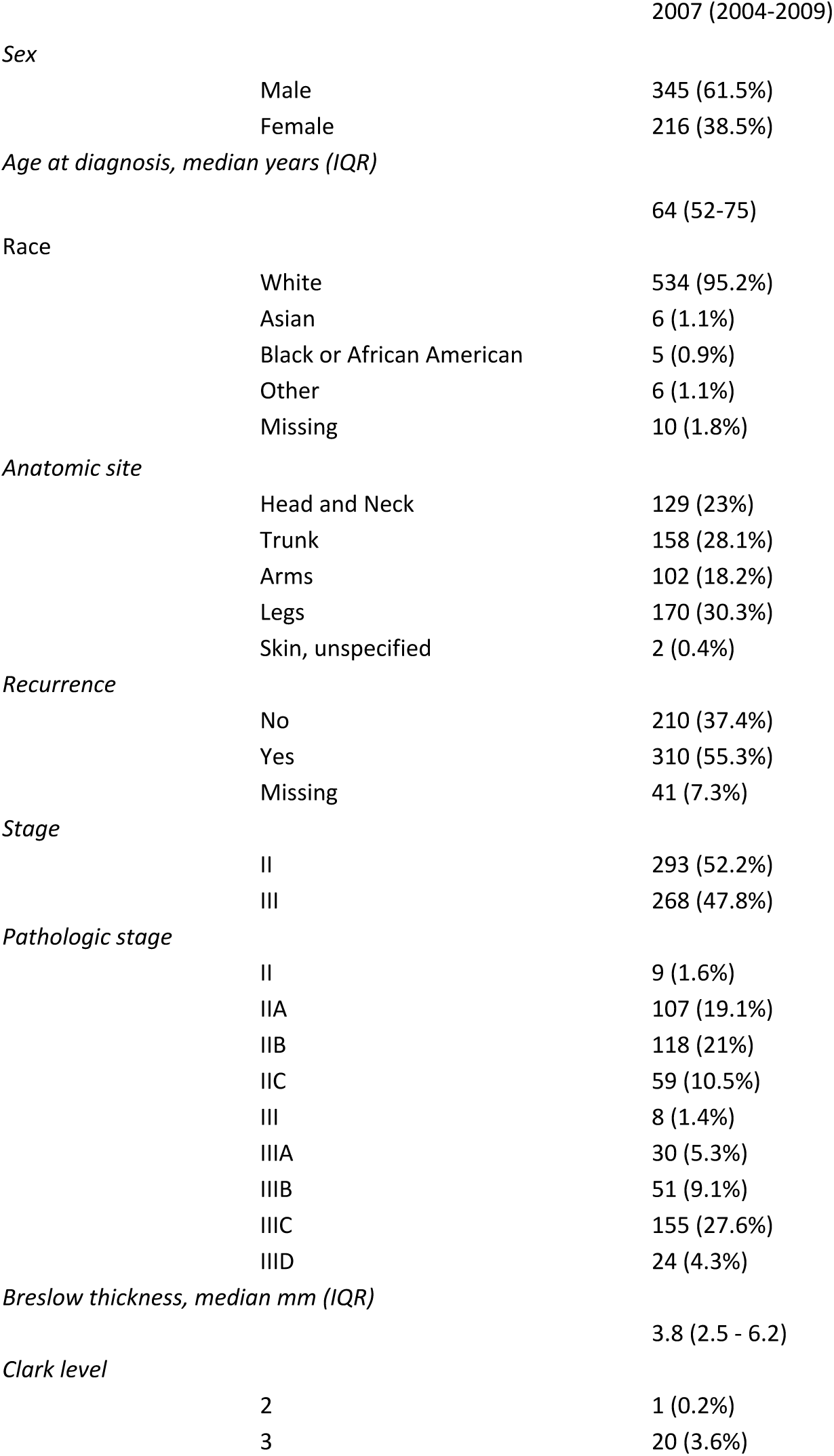
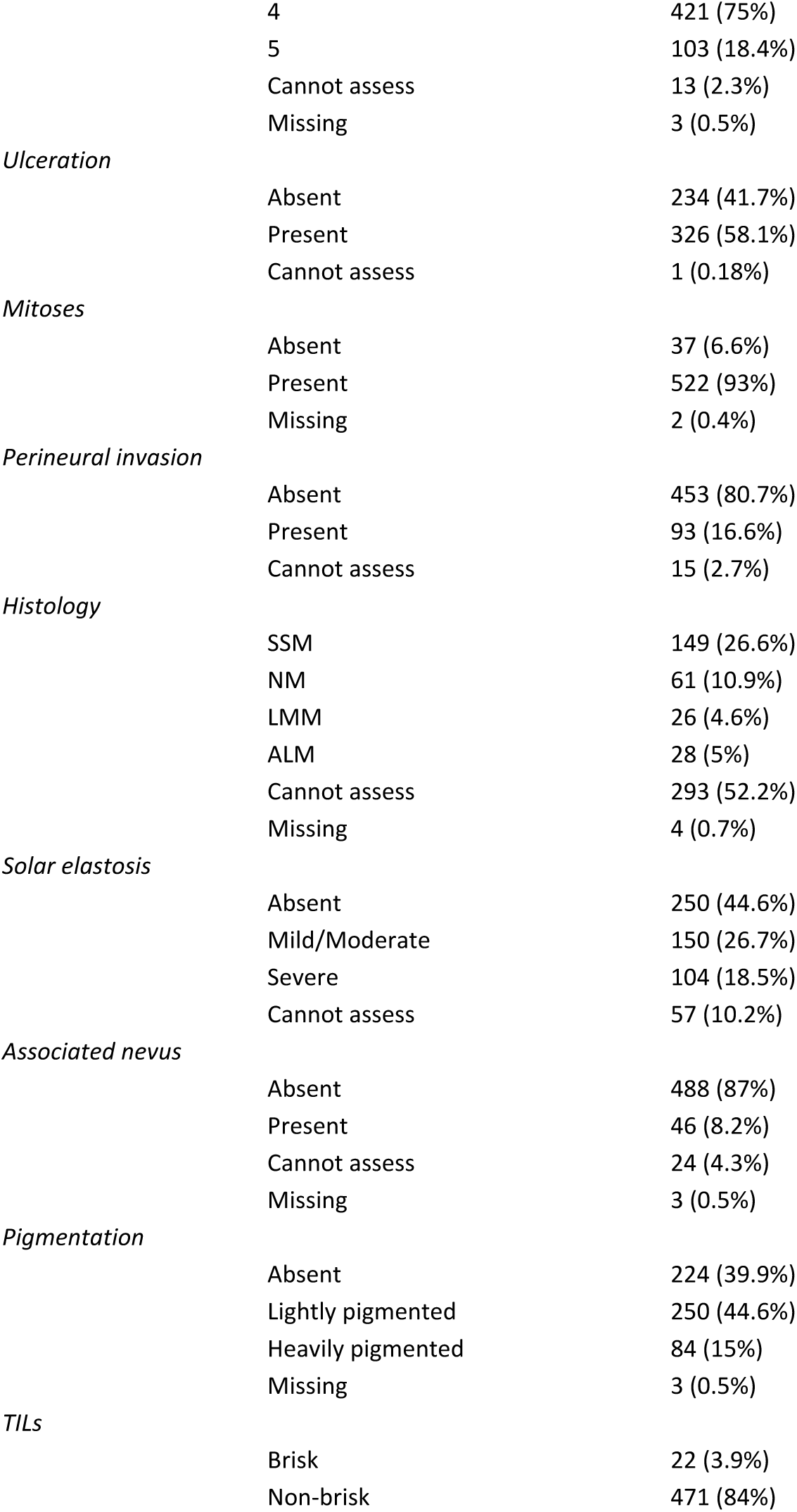
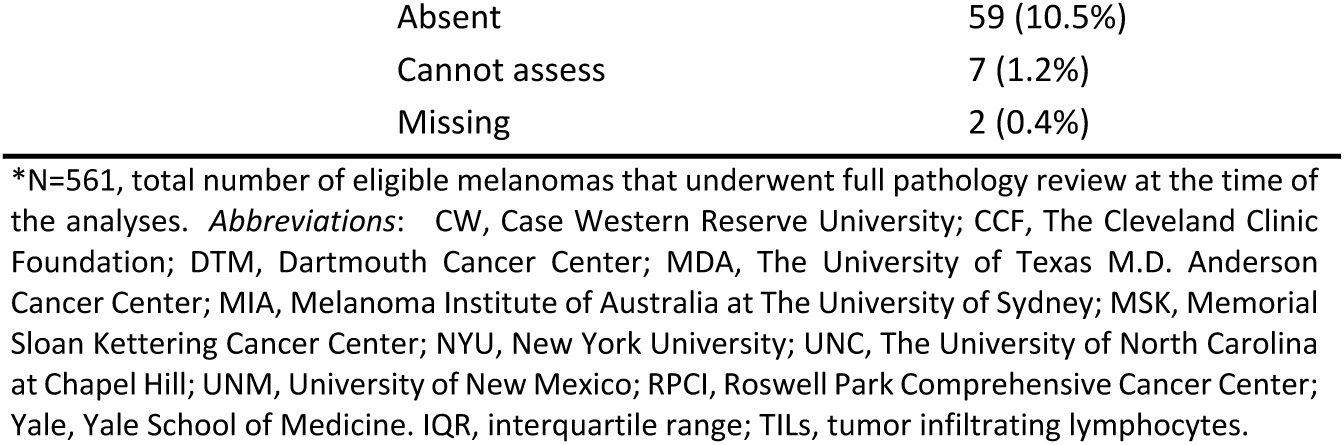
Demographic, clinical, and detailed pathologic characteristics for patients and melanomas at the time of initial diagnosis (total, n=561*).

| Patient and tumor characteristics |  | N (proportion) |
| --- | --- | --- |
| *Survival group* |  |  |
|  | Died within 5 years of diagnosis | 234 (41.7%) |
|  | Lived more than 5 years after diagnosis | 327 (58.3%) |
| *Center* |  |  |
|  | CCF | 87 (15.53%) |
|  | CW | 15 (2.7%) |
|  | DTM | 77 (13.7%) |
|  | MDA | 30 (5.3%) |
|  | MIA | 237 (42.2%) |
|  | MSK | 6 (1.1%) |
|  | NYU | 37 (6.6%) |
|  | UNC | 49 (8.7%) |
|  | RPCI | 1 (0.2%) |
|  | Yale | 22 (3.9%) |
| *Year of diagnosis, median years (IQR)* |  |  |
|  |  | 2007 (2004-2009) |
| *Sex* |  |  |
|  | Male | 345 (61.5%) |
|  | Female | 216 (38.5%) |
| *Age at diagnosis, median years (IQR)* |  |  |
|  |  | 64 (52-75) |
| *Race* |  |  |
|  | White | 534 (95.2%) |
|  | Asian | 6 (1.1%) |
|  | Black or African American | 5 (0.9%) |
|  | Other | 6 (1.1%) |
|  | Missing | 10 (1.8%) |
| *Anatomic site* |  |  |
|  | Head and Neck | 129 (23%) |
|  | Trunk | 158 (28.1%) |
|  | Arms | 102 (18.2%) |
|  | Legs | 170 (30.3%) |
|  | Skin, unspecified | 2 (0.4%) |
| *Recurrence* |  |  |
|  | No | 210 (37.4%) |
|  | Yes | 310 (55.3%) |
|  | Missing | 41 (7.3%) |
| *Stage* |  |  |
|  | II | 293 (52.2%) |
|  | III | 268 (47.8%) |
| *Pathologic stage* |  |  |
|  | II | 9 (1.6%) |
|  | IIA | 107 (19.1%) |
|  | IIB | 118 (21%) |
|  | IIC | 59 (10.5%) |
|  | III | 8 (1.4%) |
|  | IIIA | 30 (5.3%) |
|  | IIIB | 51 (9.1%) |
|  | IIIC | 155 (27.6%) |
|  | IIID | 24 (4.3%) |
| *Breslow thickness, median mm (IQR)* |  |  |
|  |  | 3.8 (2.5 - 6.2) |
| *Clark level* |  |  |
|  | 2 | 1 (0.2%) |
|  | 3 | 20 (3.6%) |
|  | 4 | 421 (75%) |
|  | 5 | 103 (18.4%) |
|  | Cannot assess | 13 (2.3%) |
|  | Missing | 3 (0.5%) |
| *Ulceration* |  |  |
|  | Absent | 234 (41.7%) |
|  | Present | 326 (58.1%) |
|  | Cannot assess | 1 (0.18%) |
| *Mitoses* |  |  |
|  | Absent | 37 (6.6%) |
|  | Present | 522 (93%) |
|  | Missing | 2 (0.4%) |
| *Perineural invasion* |  |  |
|  | Absent | 453 (80.7%) |
|  | Present | 93 (16.6%) |
|  | Cannot assess | 15 (2.7%) |
| *Histology* |  |  |
|  | SSM | 149 (26.6%) |
|  | NM | 61 (10.9%) |
|  | LMM | 26 (4.6%) |
|  | ALM | 28 (5%) |
|  | Cannot assess | 293 (52.2%) |
|  | Missing | 4 (0.7%) |
| *Solar elastosis* |  |  |
|  | Absent | 250 (44.6%) |
|  | Mild/Moderate | 150 (26.7%) |
|  | Severe | 104 (18.5%) |
|  | Cannot assess | 57 (10.2%) |
| *Associated nevus* |  |  |
|  | Absent | 488 (87%) |
|  | Present | 46 (8.2%) |
|  | Cannot assess | 24 (4.3%) |
|  | Missing | 3 (0.5%) |
| *Pigmentation* |  |  |
|  | Absent | 224 (39.9%) |
|  | Lightly pigmented | 250 (44.6%) |
|  | Heavily pigmented | 84 (15%) |
|  | Missing | 3 (0.5%) |
| *TILs* |  |  |
|  | Brisk | 22 (3.9%) |
|  | Non-brisk | 471 (84%) |

|  |  |
| --- | --- |
| Absent | 59 (10.5%) |
| Cannot assess | 7 (1.2%) |
| Missing | 2 (0.4%) |

\*N=561, total number of eligible melanomas that underwent full pathology review at the time of the analyses. *Abbreviations:* CW, Case Western Reserve University; CCF, The Cleveland Clinic Foundation; DTM, Dartmouth Cancer Center; MDA, The University of Texas M.D. Anderson Cancer Center; MIA, Melanoma Institute of Australia at The University of Sydney; MSK, Memorial Sloan Kettering Cancer Center; NYU, New York University; UNC, The University of North Carolina at Chapel Hill; UNM, University of New Mexico; RPCI, Roswell Park Comprehensive Cancer Center; Yale, Yale School of Medicine. IQR, interquartile range; TILs, tumor infiltrating lymphocytes.

### Co-extracted tumor RNA and DNA, and gDNA

Results from the preliminary assessment of two co-extraction kits done through an independent, pilot set of FFPE tissues include quantity and quality assessment, as well as genetic and epigenetic testing (S2 Appendix, S1 Fig, S2 Fig). For the current report and ongoing study, the characteristics of the FFPE target tissue included in the RNA-DNA co-extraction (tumor FFPE tissue) and in the DNA extraction (non-tumor FFPE tissue) are shown on Table 2. The total amount of RNA and dsDNA obtained per case increased with the size of the target tumor tissue, but this was not the case for the dsDNA obtained from the target non-tumor FFPE (S3 Fig). Quantity and quality characteristics of tumor and non-tumor DNA, and tumor RNA, are shown on Table 3. Inter-batch replicas had very similar miRNA expression profiles (correlation coefficient r>0.9), indicating stability of the miRNA in the sample. In general, no significant differences were noticed across batches of extracted tumors, or lots of reagents. Fingerprinting, testing with SampleID (Agena Bioscience), and/or NGS revealed six unresolvable discrepant cases for which new specimens were requested. In four of these, we found differences between the genetic and reported sex, and between tumor and paired non-tumor DNAs. Germline DNAs were extracted from 375 FFPE non-tumor tissues (279 sets of sections and 96 sets of curls), 15 blood samples, and one saliva sample.

**Table 2.** Assessment of target FFPE tissue areas for extractions of nucleic acids from tumor and non-tumor specimens.

|  | Tumor, N=685<sup>^</sup> | Non-tumor, N=279<sup>^*</sup> |
| --- | --- | --- |
| Number of sections, median (range) | 7 (4-25) | 5 (1-15) |
| % tumor, median (range) | 80 (15-95) | 0 (0-40) |
| Pigmentation visible, N | 44 | 13 |
| Sections thickness, mm median (range) | 10 (5-10) | 10 (10-10) |
| Length, mm median (range) | 10.5 (2-98) | 18 (1-46) |
| Width, mm median (range) | 3 (0.5-21) | 6 (0.25-23) |
| Area per section, mm<sup>2</sup> median (range) | 36 (1.8-480) | 98 (1-600) |
| Total tissue area mm<sup>2</sup> median (range) | 273 (12.6-4950) | 510 (5-4725) |
| *Time elapsed, days (sectioning to extraction) median (range) | 163 (15-655) | 112(30-663) |

<sup>^</sup>\*Number of tissues extracted by the March 7th, 2021 cut-off (this report). \*Counts do not include 'curls' or tissue sections placed in tubes (n=96), saliva (n=1), or blood/buffy coat (n=209). Note that characteristics are for tumors which underwent initial central histopathology review. The targeted tissue refers to the histopathology-guided and marked tissue areas, deemed suitable for scraping and extraction of nucleic acids.

**Table 3.**
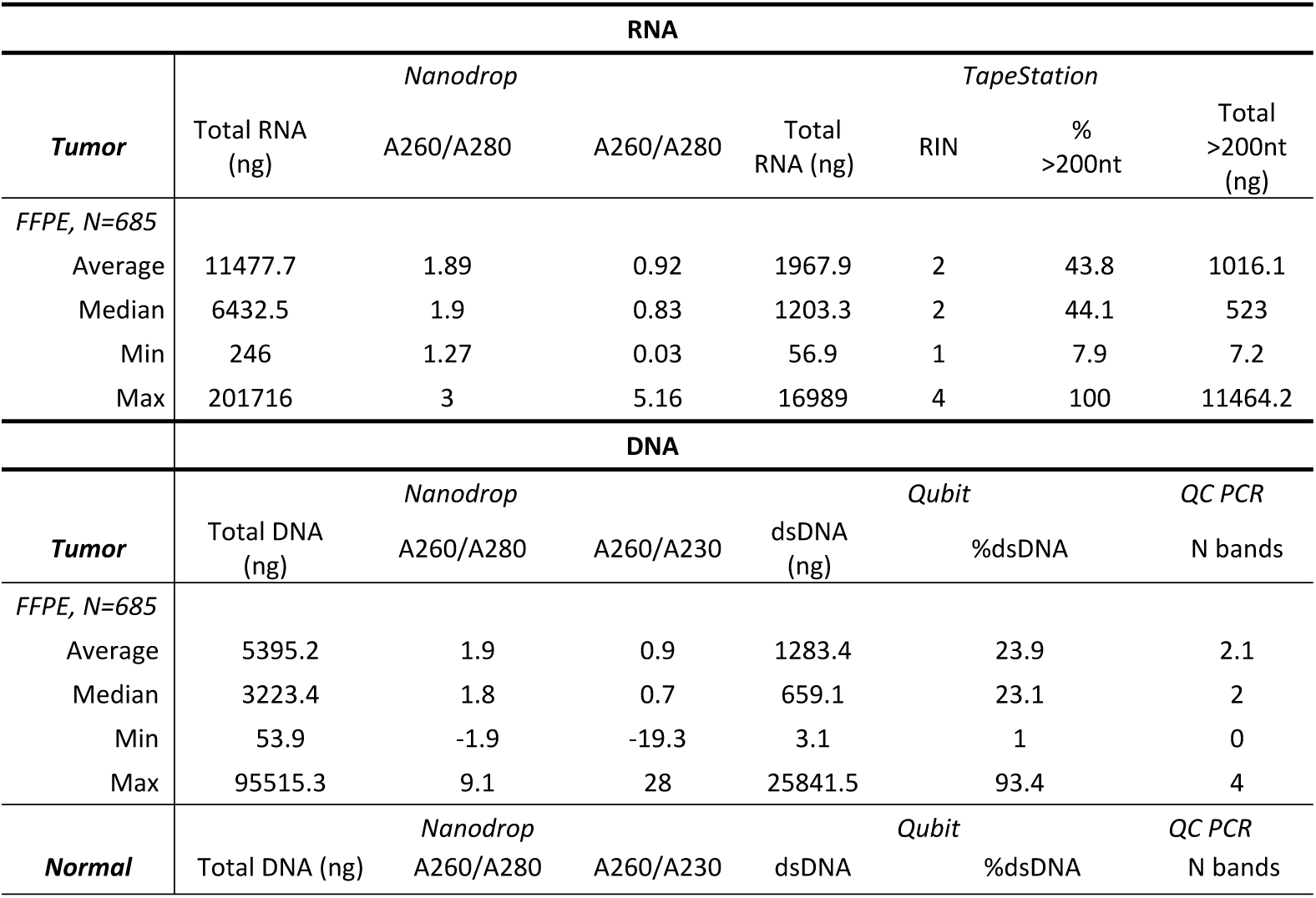
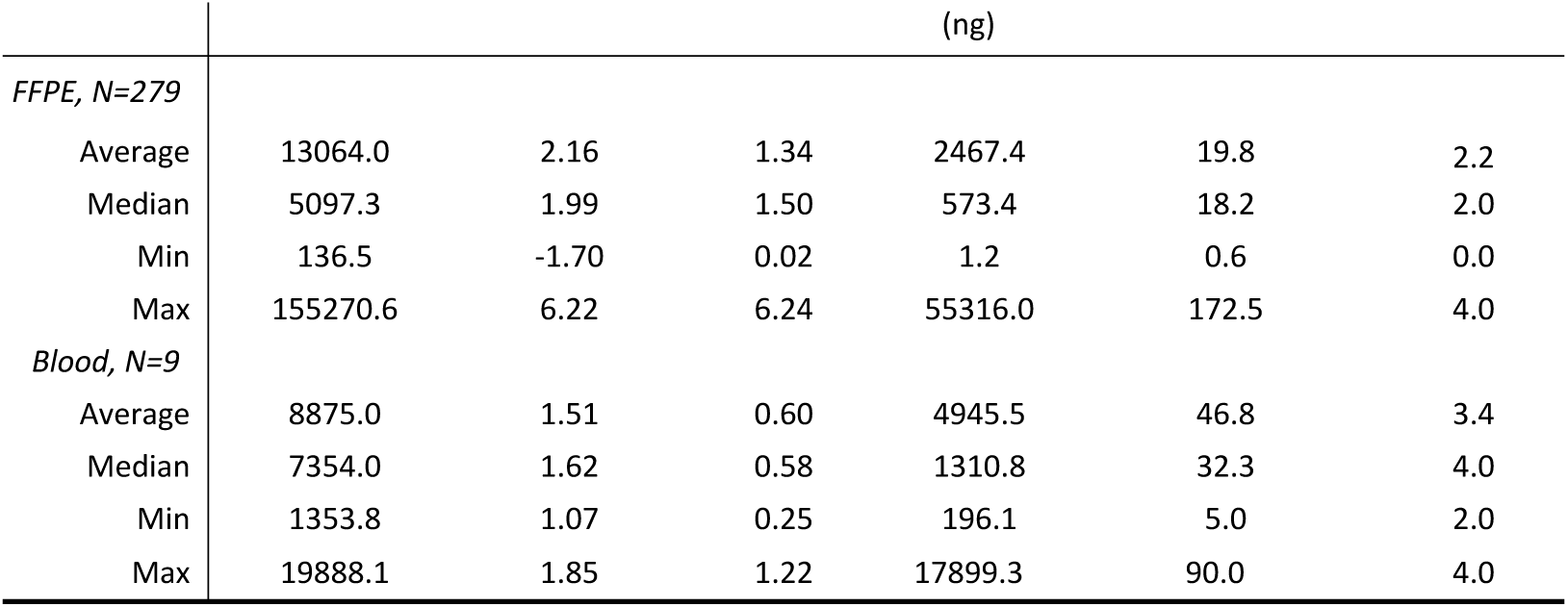
Characteristics of RNA and DNA obtained from tumor and non-tumor tissues.

### Melanoma characteristics and quantity/quality of Nucleic Acids (NA)

The tumor characteristics significantly associated with the obtained NA quantity and/or quality are depicted on Fig 2 and S4 Fig. There was a positive correlation between the extracted target tissue area and the obtained DNA (Fig 2A) and RNA yield (Fig 2B). Greater quantities of RNA relative to the extracted tumor area were obtained among stage III melanomas compared to those with stage II: 39.5 vs 32.4 ng/mm2 RNA (p<0.01); but for DNA this difference did not reach statistical significance (4.6 vs 3.9 ng/mm2 dsDNA, p=0.06). No associations were observed between the quantity of co-extracted DNA/RNA and tumor Breslow thickness, tumor purity (% tumor cells), or time elapsed between tissue block sectioning to co-extraction. Time elapsed, however, had an inverse statistically significant association with amplicon size on QC-PCR (Fig 2C). TILs were significantly associated with greater amounts of dsDNA (Fig 2D) and purity (S4A Fig). Presence of TILs was significantly associated with the ability to generate amplicons >200bp (S4B Fig). There was a positive association between pigmentation and quantities of total RNA obtained, with heavily pigmented tumors rendering higher yields compared to both lightly pigmented and non-pigmented tumors (Fig 2E). DV200 values were also higher in RNAs extracted from pigmented lesions (Fig 2F). Ulceration was significantly associated with greater DNA A260/A280 (Fig 2G) and RNA A260/A280 values (p<0.045, not shown). DNA and RNA samples obtained from heavily pigmented tumors had lower A260/A280 ratios (Fig 2H-I). DNA from pigmented tumors amplified shorter fragments (S4C Fig). Stage III, tumor purity, and presence of ulceration were all positively associated with DNA A260/A230 ratios (p=0.04, p=0.04, and p=0.004, respectively, data not shown). Evaluation of absorbance (260/280, and 260/230) ratios was limited to samples with concentrations ≥20ng/µL as lower concentrations render inaccurate data. Fig 3 depicts the effect of case characteristics (Fig 3A), time elapsed from sectioning to co-extraction (Fig 3B), and tumor characteristics (Fig 3C-D), on the recommendations to proceed with the synthesis of libraries. In addition, we observed a statistically significant association between presence of ulceration and DNA-QC ‘failures’, with 15% of DNA samples categorized as failed among ulcerated melanomas compared to 28% failed DNAs among those without ulceration (p<0.001, data not shown). The effect of DNA and RNA quality characteristics (purity, fragmentation/integrity) on the recommendations to proceed with (a) the synthesis of libraries and (b) MSK-IMPACT™ assay are shown in S5 Fig.

**Fig 2.**
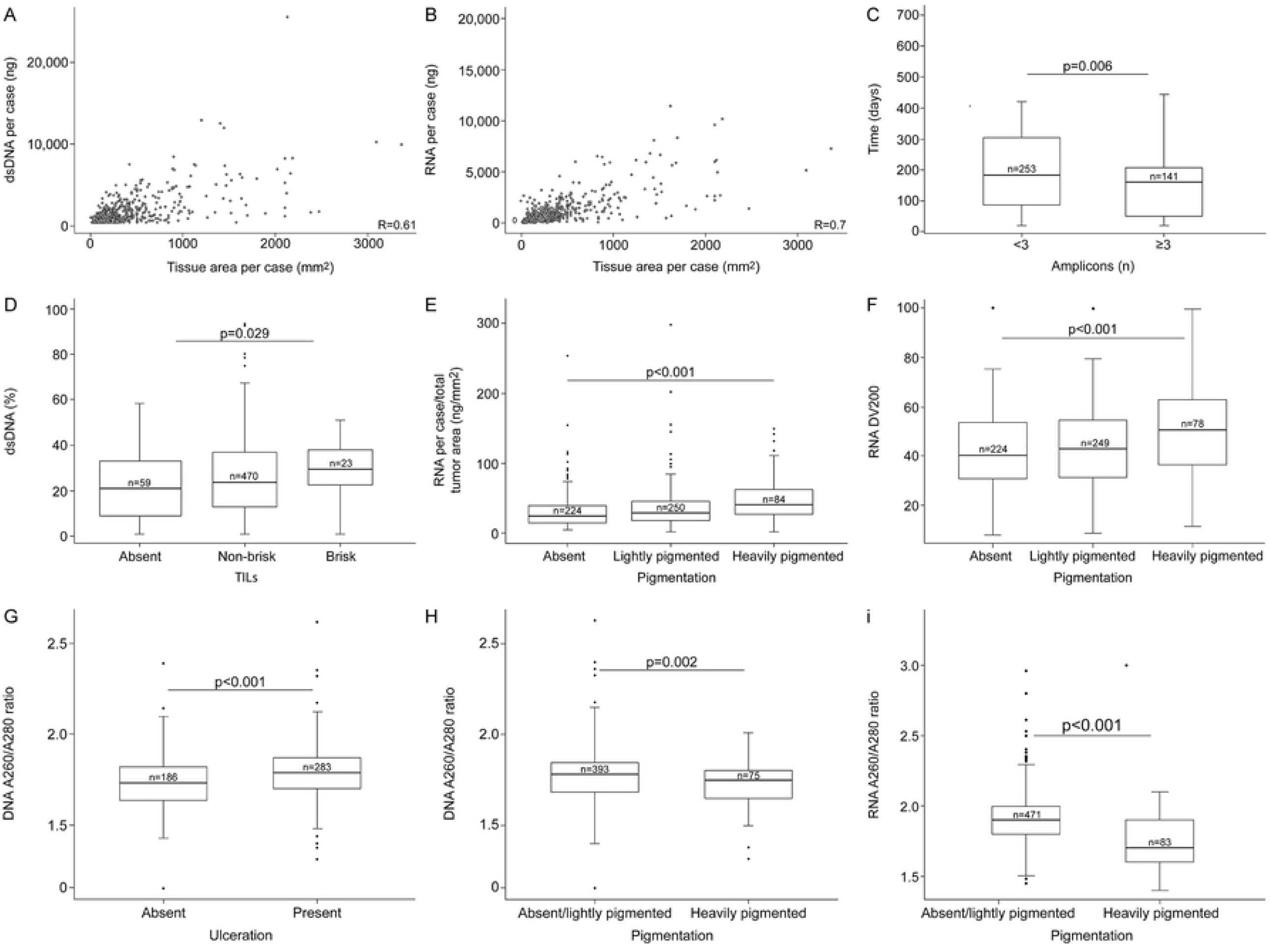
Effect of melanoma characteristics and time elapsed between tissue sectioning and co-extraction, on the quantity and quality of DNA/RNA.

**Fig 3.**
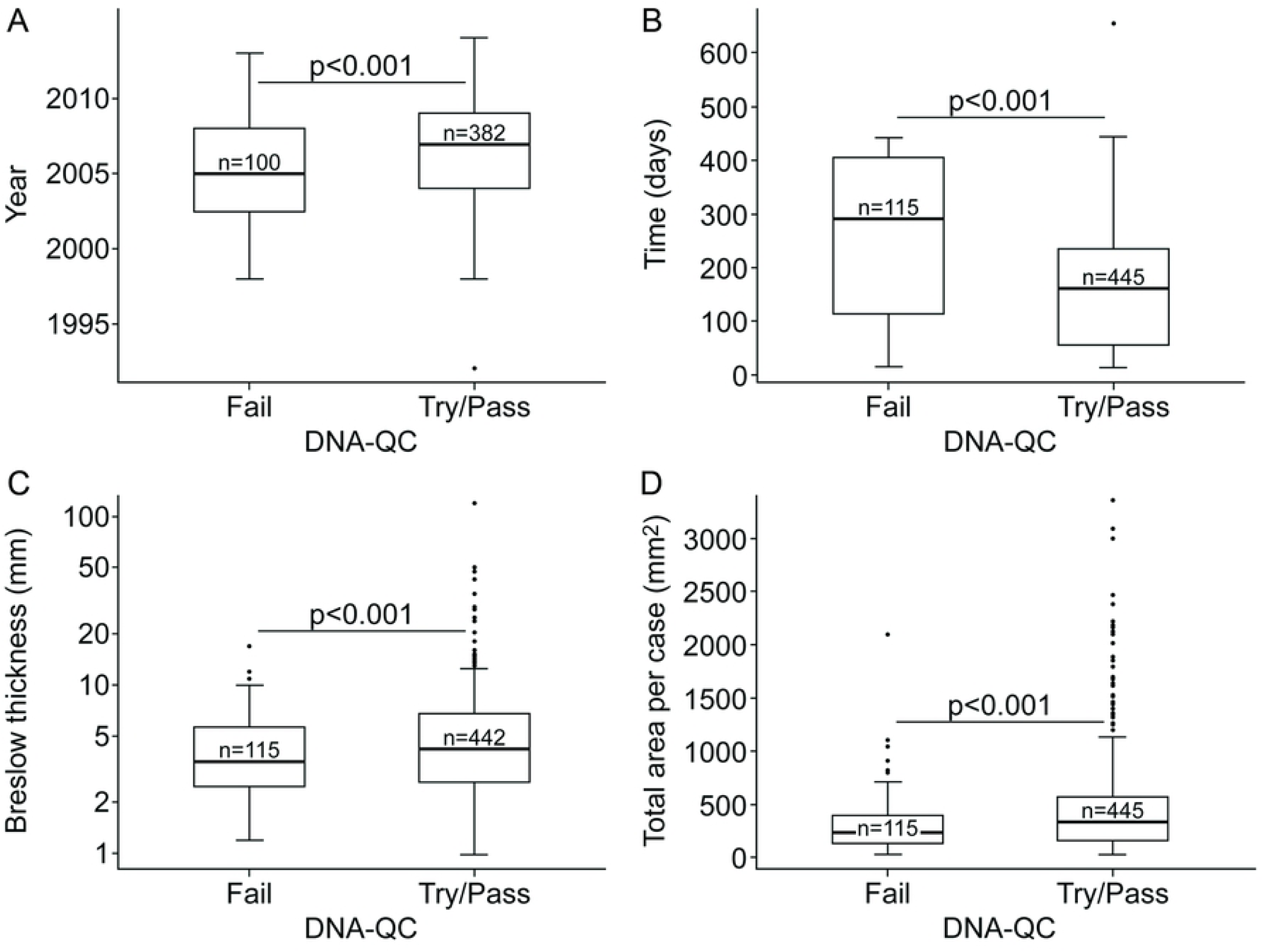
Effect of case and tumor characteristics, and time elapsed between sectioning and co-extraction on the core’s DNA-QC score.

### Distribution of nucleic acids for genetics and epigenetics testing, and preliminary QC

Almost all RNA samples, 683/685 (99%), qualified for distribution for miRNA expression analysis with the Nanostring nCounter® Human v3 miRNA Expression assay, 467 (68%) DNA samples for methylation profiling with the Illumina Infinium MethylationEPIC arrays, and 560 (82%) DNA samples, for somatic mutation profiling by NGS with the MSK-IMPACT™ assay (Fig 4). In 446 (65%) cases, aliquots of RNA and DNA were distributed for testing with all three platforms. We compared the melanoma and tissue characteristics among cases with samples that qualified across testing platforms to evaluate potential biases or imbalances, and these characteristics are shown on Table 4.

**Fig 4.**
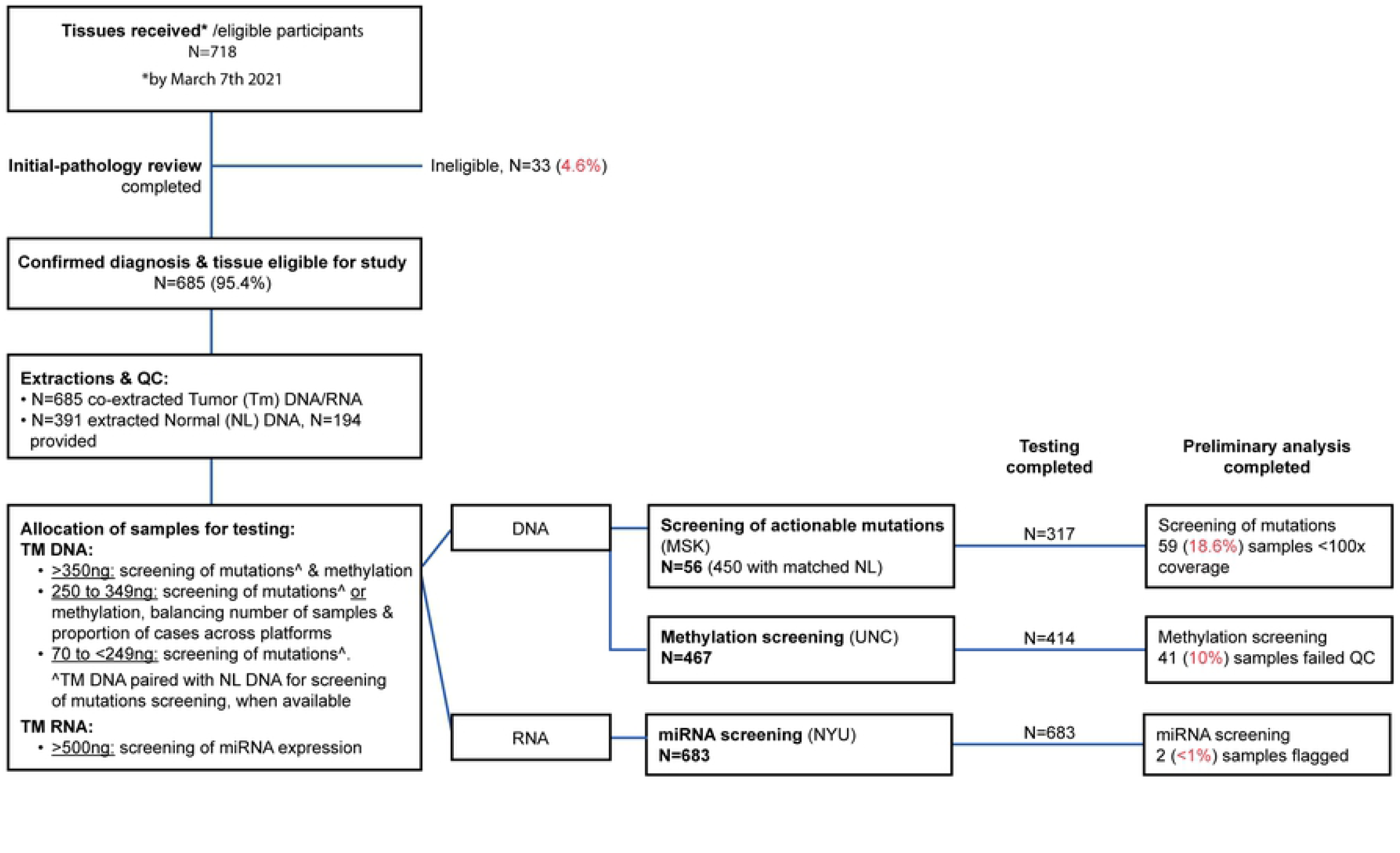
Biospecimens received, processed, and distributed for testing.

**Table 4.** **Characteristics of participants and their melanomas for cases and controls with available full histopathology review and with samples allocated for testing, by miRNA, methylation, and mutation profiling platforms.**

|  | All samples | Tested on all 3 platforms* | miRNA* | methylation* | NGS mutations* |
| --- | --- | --- | --- | --- | --- |
| *N* | 561 | 396 | 559 | 414 | 482 |
| *Age* |  |  |  |  |  |
| median years (IQR) | 64 (52-75) | 64 (54-75) | 64 (52-75) | 64 (53-75) | 64 (53-76) |
| *Sex* |  |  |  |  |  |
| male | 345 (61.5%) | 248 (62.6%) | 343 (61.4%) | 262 (63.3%) | 299 (62.0%) |
| female | 216 (38.5%) | 148 (37.4%) | 216 (38.6%) | 152 (36.7%) | 183 (38.0%) |
| *Survival Group* |  |  |  |  |  |
| survived ≤5 years | 327 (58.3%) | 212 (53.5%) | 327 (58.5%) | 225 (54.4%) | 271 (56.2%) |
| died <5 years | 234 (41.7%) | 184 (46.5%) | 232 (41.5%) | 189 (45.6%) | 211 (43.8%) |
| *Breslow thickness* |  |  |  |  |  |
| median mm (IQR) | 3.8 (2.5-6.2) | 4.2 (2.8-6.8) | 3.8 (2.5-6.2) | 4.2 (2.7-6.8) | 4.0 (2.5-6.5) |
| not available | 3 | 2 | 2 | 2 | 2 |
| *Ulceration* |  |  |  |  |  |
| present | 326 (58.2%) | 248 (62.8%) | 325 (58.2%) | 258 (62.5%) | 289 (60.0%) |
| absent | 234 (41.8%) | 147 (37.2%) | 233 (41.8%) | 155 (37.5%) | 192 (40.0%) |
| *not available* | 1 | 1 | 1 | 1 | 1 |
| *Mitoses* |  |  |  |  |  |
| present | 522 (93.4%) | 377 (95.2%) | 522 (93.4%) | 392 (94.7%) | 453 (94.0%) |
| absent | 37 (6.6%) | 19 (4.8%) | 37 (6.6%) | 22 (5.3%) | 29 (6.0%) |
| *not available* | 2 |  |  |  |  |
| *Pigmentation* |  |  |  |  |  |
| absent/ not pigmented | 224 (40.1%) | 160 (40.9%) | 223 (40.1%) | 169 (41.0%) | 193 (40.2%) |
| lightly pigmented | 250 (44.8%) | 174 (43.9%) | 249 (44.8%) | 179 (43.5%) | 214 (44.6%) |
| heavily pigmented | 84 (15.1%) | 60 (15.2%) | 84 (15.1%) | 64 (15.5%) | 73 (15.2%) |
| *not available* | 3 | 2 | 3 | 2 | 2 |
| absent/lightly pigmented | 474 (85.0%) | 334 (84.8%) | 472 (84.5%) | 348 (84.5%) | 407 (85.8%) |
| heavily pigmented | 84 (15.0%) | 60 (15.2%) | 84 (15.1%) | 64 (15.5%) | 73 (15.2%) |
| *not available* | 3 | 2 | 3 | 2 | 2 |
| *Stage* |  |  |  |  |  |
| II | 293 (52.2%) | 200 (50.5%) | 292 (52.2%) | 207 (50.0%) | 252 (52.5%) |
| III | 268 (47.8%) | 196 (49.5%) | 267 (47.8%) | 207 (50.0%) | 230 (47.7%) |

\*N indicate availability of samples, data and data-analysis by the cut-off date. Allocation of samples for testing depends on the availability of tumor RNA (miRNA expression), tumor DNA (methylation), tumor and germline DNA (NGS, next generation sequencing for somatic mutations). For limited DNA, distribution for methylation or NGS contemplates survival group and overall balanced numbers across these two platforms. Proportions within each column sum up to 100% and do not consider the few unavailable datapoints, as shown above. Age, tumor Breslow thickness, and stage correspond to the initial diagnosis. Breslow and ulceration correspond to the maximum value reported either at diagnosis or during the centralized pathology review.

Upon testing, 41 of 414 (10%) DNA samples with available methylation data for the present study failed QC due to low intensity probes (p>0.05) or insufficient single-sample Noob-normalization or BMIQ-normalization. We evaluated the effect of survival, marginally associated with failures (Fig 5A), to assess differences in the attrition between cases and controls. Characteristics associated with methylation screening failure were year of diagnosis (i.e. age of the blocks) (Fig 5B), time elapsed from sectioning to extraction (Fig 5C), %dsDNA (Fig 5D), and amplifiability (Fig 5D). Six of 683 (1%) RNA samples failed the Nanostring-based QC. These samples were flagged and then deemed unsuitable for data analysis because of their low proportion of probes above the minimal threshold. Inter-batch replicas showed a very similar miRNA expression profile before (r>0.8) and after correcting for batch effect (r>0.9). There were no variables found in association with Nanostring flags or failures.

**Fig 5.**
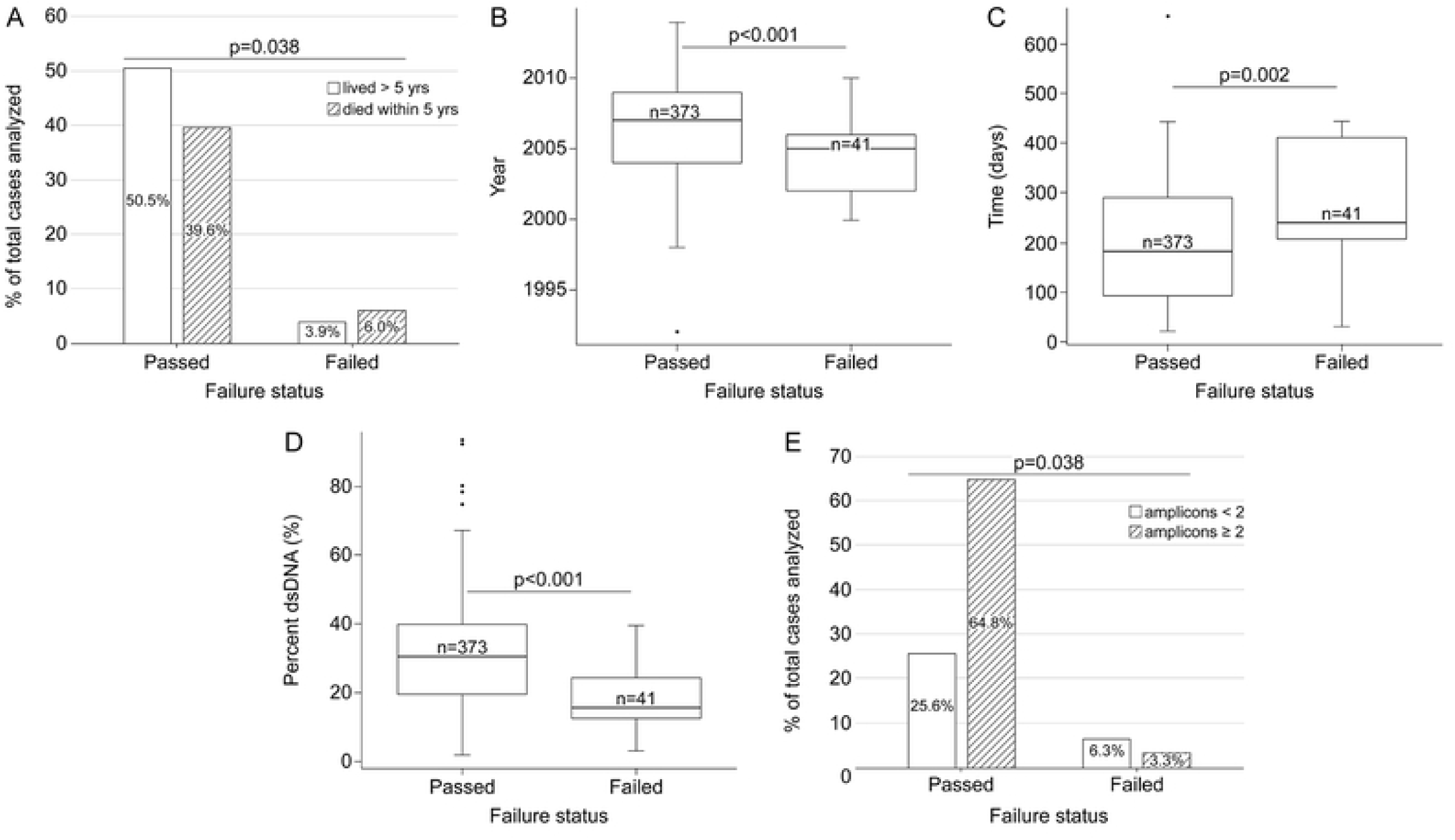
Effect of case, tumor, DNA characteristics, and logistics, on the success of methylation screening.

Of the samples sent for mutations screening with the MSK-IMPACT™ assay, 5 samples were subsequently deemed ineligible, 317/560 (57%) were run through the pipeline, and had data available for the present analysis. On average, the coverage was 249x, with a range of 22x to 762x, and 59 (18.6%) samples had coverage below 100x (Fig 4). The recommendations to proceed (or hold samples back) with the synthesis of libraries or MSK-IMPACT assay were significantly associated with coverage (p<0.01) (S6 Fig); however, no significant associations were found between the amount of tissue extracted or any of the NA characteristics with samples scored as ‘fail’ by the genetics core (DNA-QC, Lib-QC). For details on data availability, see S1 Appendix.

## Discussion

We have delineated a step-by-step approach for the handling of biospecimens and for data collection in a large multi-center international study, including quality assurance and quality control, in preparation for multi-omics testing from small sample size archival primary melanoma tissue. A unique aspect of this study is the testing of DNA and RNA co-extracted from the same cell lysate obtained from FFPE tissues. This co-extraction is critical for the proper integration of data obtained from multi-omics platforms utilized for the screening of mutations, methylation, and miRNA profiling, with protein expression and mRNA planned in the near future.

FFPE tissue blocks of primary melanomas are extremely valuable as they represent the target lesion or ‘index tumor’ of interest and are accompanied by a vast array of clinicopathologic information. The process of formalin fixation and paraffin embedding helps conserve the cells topography, and blocks can be stored for decades without the need of ultra-low temperatures storage. However, it is well known that greater amounts and quality of nucleic acids may be obtained from fresh frozen tissues. Melanomas in particular, compared to other solid tumors, are most often very small at the time of diagnosis, and for purposes of accurate diagnosis the whole biopsy needs to be FFPE-processed. Thus, once diagnosis is completed as part of the standard care, only a limited amount of FFPE tissue remains available for research. While archived FFPE tissues are more likely to be available and more convenient for precision medicine, they present some challenges (18). Because formalin forms cross-links during fixation that cause fragmentation during DNA extraction, and introduces contaminants, the use of FFPE can introduce artifacts in molecular profiling, although newer laboratory techniques, and analytical tools, help reduce the errors.

Our priority is obtaining sufficient ‘quality’ DNA and RNA without exhausting FFPE blocks. In most cases, tumor tissues were procured as per our standard operating procedures, and when necessary fewer or greater number of sections were provided when necessary (due to scarcity of tissue or evidence of particularly small tumor areas). Non-tumor FFPE tissues, characterized by a lower cell (and nuclei) density, yield lower and less predictable amounts of DNA. Few centers have access to blood, buccal cells/saliva, or other tissue sources of non-tumor germline DNA. While surrounding tumor tissue may have some genetic change due to field effects, it is still preferable to utilize adjacent normal tissue when no other source of germline DNA is available. We preferentially distribute tumor DNA paired with matched normal for somatic mutations profiling with MSK-IMPACT™; however, matched normal samples are not always available, or sufficient. While matched tumor-normal DNAs doubles the NGS costs, this approach provides ‘clean’ or accurate somatic mutation calling, free of germline artifacts. In addition, germline DNA allows the uncovering of potential mismatches, discrepancies, or contamination, through DNA fingerprinting. Of note, in this study we uncovered a very small fraction of discrepant samples, and even fewer remained unsolvable (0.9%) after rematching, or replacement of germline DNA. For melanomas in which the matching germline DNA is not available or the amount and/or quality inadequate, we submit tumor-only batches for analyses against a pool of normal DNA. To this effect, a robust pipeline has been developed to identify germline artifacts, taking into account coverage, allele level, copy number, and tumor purity (Arshi Arora, manuscript in preparation). A related concern is the choice of volume for the elution of nucleic acids during the extraction (normal tissue) and co-extraction (tumors). On the one hand, greater eluates recover more DNA from the column but at lower concentrations, risking greater inaccuracy. On the other hand, with eluates that are too small, more DNA is lost to the column, capillary action, and to quantity and quality measurements. The ideal volume needs to be adapted according to preliminary co-extractions on similar tissues. Samples <20ng/ul are less reliable for determining purity accurately and this, plus much more stringent criteria, may contribute to the differences observed in the QC assessments obtained by our InterMEL Biocore for the NGS pipeline versus the CLIA-approved process that was developed by MSK for clinical use.

In ideal conditions, tissues would render sufficient nucleic acids for QC and distribution to all three testing platforms. In our experience with small primary melanomas, 35% of the eligible cases had insufficient material for all 3 omics platforms: somatic mutations, methylation, and miRNA. Individually, 99.7% of the eligible cases had sufficient RNA for miRNA analysis, 78.8% for NGS, and 68.2% for methylation. To evaluate methylation, samples need to undergo an initial treatment with bisulfite, to convert unmethylated Cs to Ts. This treatment converts double-stranded DNA into more labile and unpaired single stranded DNA. Older tissue blocks and sections with more fragmented co-extracted DNA, will be naturally more susceptible to degradation, resulting in greater attrition, despite the use of a kit to restore degraded FFPE DNA (Fig 5). Melanomas with double-stranded DNA quantities below the threshold for methylation and mutation screening (∼320 to 350ng double-stranded DNA), are adjudicated in nearly equal numbers for testing with one or the other DNA platform. In addition, in making this choice we considered their case-control status as well as availability (or not) of germline DNA for NGS. We also evaluated whether certain patient and tumor characteristics biased the distribution of samples across testing platforms. Age and sex were comparable across testing platforms, while some differences were noticed in the proportion of samples from patients who were stage III, died of melanoma within five-years, had thicker tumors, or with ulceration or mitoses (Table 4). The differences were not large, and these clinicopathologic variables will be included as covariates in downstream/formal statistical analyses. Another important consideration is the timing of tissue sectioning. Because centralized block sectioning is not feasible, and most contributing centers are geographically distant from MSK, we wanted to minimize the time elapsed between sectioning and nucleic acid co-extraction, which is fully dependent on diagnostic confirmation/ histopathology review and marking of H&Es. The COVID-19 pandemic added unanticipated delays through complete closures in the first couple of months, followed by a few months of limited access to the research facilities, and major delays in the supply-chain, with sectioned tissues awaiting extractions for a very prolonged time. Indeed, the elapsed time had a detrimental effect on the DNA performance, which was evident through the ability to amplify fragments ≥300nt, synthesis of libraries for NGS, and the success of the methylation screening (Figs 2, 3; and 5).

Interestingly, we obtained greater amounts of RNA in heavily pigmented tissue. We posit that the melanin content decreases the pH, and that this may inhibit the enzymatic activity of the RNAses, otherwise ubiquitously present, an idea further supported by the greater proportion of RNA strands >200nt in heavily pigmented melanomas (Fig 2). Melanin is known to inhibit some enzymatic reactions (19, 20) including amplification with Taq polymerase. Not surprisingly, samples obtained from heavily pigmented melanomas, on average, amplified shorter fragments (S4 Fig); however, we chose to not add an extra treatment to the samples as this would have affected the total yield. Unlike pigmentation, TILs and ulceration improved the DNA A260/A280 ratios, and samples from melanomas with brisk TILs showed greater %dsDNA and amplifiability, perhaps due to more actively dividing cells (21) resulting in increased cellularity. Samples with greater purity and %dsDNA were better scored by the core, but not those with higher RNA integrity. We have not found an explanation for this effect yet, although, no single variable or set of DNA variables was associated alone with the core’s prediction for failure, or with the coverage obtained through the MSK-IMPACT™ assay. This differs to some extend to findings from others who reported block age as the most important variable that influenced sample viability for amplicon-based library construction (22). For methylation, on the other hand, there are clear associations between year of diagnosis (as a proxy of block’s age), time elapsed between sectioning and extraction, %dsDNA, and amplifiability, with failures (Fig 5).

Another important consideration is the jurisdiction of the centers providing tissues and data. For instance, in 2020, a European center expressed interest in joining our efforts and contributing with a substantial number of cases. However, to comply with the General Data Protection Regulation 2016/679 (regulation in EU law, on data protection/privacy in the EU), a thorough analysis, numerous audits and reviews are taking place in relation to detailed information on (i) where specimens and data from EU will be located, (ii) who will have access to samples and data, (iii) how data and specimens are protected, and (iv) which systems and applications will be used to process, store, and run data derived for the samples from EU. This major delay may potentially affect the feasibility of including these specimens if samples cannot be procured and tested in time for analyses, especially since tissues cannot be sectioned in advance. International collaborations are important for ensuring worldwide representation of patient samples. Investigators should plan for these international regulatory steps, when planning future consortia.

In summary, we find only two variables that, to some extent, could be controlled in future studies of archival tumors and that have a detrimental effect on the quality of the extracted DNA from melanoma tumors: age of the FFPE tissue blocks, and time elapsed between block sectioning and co-extraction. Because of the large individual sample variability, we were not able to define any variable predictive of success on NGS; consequently, we will continue using a combination of quantity and quality parameters when choosing and preparing aliquots for screening DNA samples with the MSK-IMPACT™, and we will continue aiming to decrease the time elapsed between sectioning, extraction, and testing. The co-extracted RNA does not appear to be affected by the age of the block or time elapsed, but surprisingly, presence of melanin might impart some protection against RNA degradation in FFPE tissues. The greater RNA amounts and RIN values obtained in pigmented melanomas, to the best of our knowledge, are not artifactual, and are possibly due to a change in acidity or pH. Although pigment has no impact on the success of the miRNA screening with Nanostring, our observations are intriguing and worthy of further consideration. Of the 685 eligible cases, 65% of the melanomas rendered sufficient material for testing by all three omics platforms. The implication is that in future studies of early-stage melanomas using archival tissues, one should aim to accrue sufficient material to allow a 35% attrition, when the goal is to obtain genetic and epigenetic profiling such as ours. Furthermore, when considering the methylation failures, the ideal target would permit 40% attrition. We anticipate that these reported practical guidelines for the procurement, preparation, quality control, and testing of co-extracted DNA and RNA for the screening of actionable somatic mutations, methylation and miRNA using high-throughput platforms, as well as our data and insight will be of value to others with their ongoing and future investigations using valuable archival yet limited tissues.

## Data Availability

The data underlying this report are subject to an embargo until December 1st, 2023. Once the embargo expires, the data will be available on the National Cancer Institute dbGaP website upon request.

## Acknowledgements

This study was conducted by the InterMEL consortium, which includes the following members: Coordinating Center, University of New Mexico, Albuquerque, NM, USA; Marianne Berwick (Principal Investigator (MPI, contact PI)), Li Luo (Investigator), Tawny W. Boyce (Data Manager), Adam Z. Reynolds (Data Analyst); University of North Carolina, Chapel Hill, NC: Nancy E. Thomas (MPI), Kathleen Conway (Investigator), Sharon N. Edmiston (Research Analyst), David W. Ollila (Surgical Oncologist and Investigator), Honglin Hao (Laboratory Specialist), Eloise Parrish (Laboratory Specialist), Paul B. Googe (Dermatopathologist), Stergios J. Moschos (Oncologist), David Corcoran (Bioinformatician), Weida Gong (Bioinformatician), Amanda Vondras (Bioinformatician), Lan Lin (Database Manager); Study centers include the following: Memorial Sloan Kettering Cancer Center, New York, NY: Ronglai Shen (PI-Project 4), Colin B. Begg (Biostatistician), Arshi Arora (Biostatistician), Venkatraman Seshan (Biostatistician), Caroline E. Kostrzewa (Assistant Research Biostatistician), Klaus J. Busam (PI-Core2), Irene Orlow (Co-PI-Core 2), Jessica M. Kenney (Research Assistant/Sr. Laboratory Specialist), Keimya D. Sadeghi (Research Assistant and Data Engineer), Kelli O’Connell (Research Biostatistician), Gbemisola Elizabeth Ilelaboye (Senior Research Technician); initial optimization assays were performed at MSK by Heta Parmar, Siok Leong, and Sergio Corrales (Research Technicians); Melanoma Institute Australia, Sydney, Australia: Richard A. Scolyer (Dermatopathologist and Site PI), Anne E. Cust (Epidemiologist and Site PI), James S. Wilmott (Scientist), Graham J. Mann (Cancer Geneticist and Former Site PI), Ping Shang (Scientist), Hazel Burke (Data Manager), Peter M. Ferguson (Pathologist), Valerie Jakrot (Research Manager); British Columbia Cancer Research Center, Vancouver, Canada: Tim K. Lee (Data Coordinator); New York University, Langone Cancer Center, New York, NY: Eva Hernando (Site PI), Iman Osman (co-Investigator), Douglas Hanniford (Instructor), Diana Argibay (Laboratory Technician), Una Moran (Data Technician), for the Genomics Technology Center: Adriana Heguy (Director), Sitharam Ramaswami (Associate Research Scientist); Baylor College of Medicine, Houston, TX: Christopher I. Amos (Advisor), Ivan P. Gorlov (PI-Core 3), Dakai Zhu (Data Analyst); Roswell Park Cancer Institute, Buffalo, NY: Marc Ernstoff (Advisor), Paul N. Bogner (Dermatopathologist); The University of Texas MD Anderson Cancer Center, Houston, TX: Jeffrey E. Lee (Site PI), Shenying Fang (Biostatistician); Dartmouth Cancer Center, Lebanon, NH: Judy R. Rees (Site PI), Shaofeng Yan (Pathologist); Case Western University, Cleveland, OH: Meg R. Gerstenblith (Site PI), Cheryl Thompson (Co-Investigator); Cleveland Clinic, Cleveland, OH: Jennifer S. Ko (Dermatopathologist and Site PI), Pauline Funchain (Co-Investigator), Peter Ngo (Dermatology Fellow); Yale University Cancer Center, New Haven, CT: Marcus Bosenberg (Site PI), Bonnie E. Gould Rothberg (Former Site PI), Gauri Panse (Dermatopathologist); Columbia University Medical School, New York, NY: Yvonne M. Saenger (Site PI), Benjamin T. Fullerton (Laboratory Technician); Huntsman Cancer Institute, Salt Lake City, UT: Sheri L. Holmen (Site PI), Howard Colman (Co-Investigator), Elise K. Brunsgaard (Clinical Fellow), David Wada (Dermatopathologist); Instituto Valencia de Oncologia, Valencia, Spain: Eduardo Nagore (Site PI), Esperanza Manrique-Silva (Dermatologist), Celia Requena (Dermatologist), Victor Traves (Pathologist), David Millan-Esteban (Post-Doctoral Fellow). Patient Advocate: Michelle Rainka.

The contribution of our colleagues at the Melanoma Institute Australia and Royal Prince Alfred Hospital is gratefully acknowledged. We also wish to acknowledge the invaluable support provided at MSK by: members of the Integrative Genomics Operation (IGO), Kety Huberman, Marisa Dunigan, Agnes Viale, PhD and Neeman Mohibullah PhD; Center for Molecular Oncology & Informatics, Nicholas Socci; members of the Department of Epidemiology & Biostatistics: Alexandra Rizzatti (Administrative Assistant), Bradley Cohen (Sr. Project Manager), Sharon Bayuga (Associate Director, Clinical Research Operations – Research & Technology Management); Marketing & Communications: Susan Weil (Design & Creative Services Production Manager).

## Supporting Information

**Appendix 1: Methods**.

**Appendix 2: Preliminary Results. Appendix 3: Supporting Figures**.

**S1 Fig. Quantity and quality assessment of RNA and DNA co-extracted from FFPE**.

**S2 Fig. Performance of RNA co-extraction kit and elution on Nanostring**^**TM**^.

**S3 Fig. Effect of FFPE tissue input on the yield of co-extracted nucleic acids**.

**S4 Fig. Effect of TILs and pigmentation on DNA purity and amplifiability**.

**S5 Fig. Effect of the DNA and RNA quality on recommendations to proceed with synthesis of libraries and NGS/MSK-IMPACT**^**TM**^.

**S6 Fig. Comparison of DNA- and library-QC recommendations with coverage obtained in melanomas screened with the MSK-IMPACT**^**TM**^ **assay**.

**Table S1. InterMEL: Participants and melanoma variables and definitions**.

## References

1. Siegel RL, Miller KD, Fuchs HE, Jemal A. Cancer statistics, 2022. CA Cancer J Clin. 2022;72(1):7–33.

2. Howlader N, Noone A, Krapcho M, Garshell J, Neyman N, Altekruse S, et al. SEER Cancer Statistics Review Bethesda, MD: National Cancer Institute; 1975-2010 [updated April 2013. Available from: http://seer.cancer.gov/csr/1975_2010/.

3. Soong SJ, Ding S, Coit D, Balch CM, Gershenwald JE, Thompson JF, et al. Predicting survival outcome of localized melanoma: an electronic prediction tool based on the AJCC Melanoma Database. Ann Surg Oncol. 2010;17(8):2006–14.

4. Macdonald JB, Dueck AC, Gray RJ, Wasif N, Swanson DL, Sekulic A, et al. Malignant melanoma in the elderly: different regional disease and poorer prognosis. J Cancer. 2011;2:538–43.

5. MacKie RM. Malignant melanoma: clinical variants and prognostic indicators. Clin Exp Dermatol. 2000;25(6):471–5.

6. Gershenwald JE, Scolyer RA, Hess KR, Sondak VK, Long GV, Ross MI, et al. Melanoma staging: Evidence-based changes in the American Joint Committee on Cancer eighth edition cancer staging manual. CA Cancer J Clin. 2017;67(6):472–92.

7. Howard MD, Wee E, Wolfe R, McLean CA, Kelly JW, Pan Y. Anatomic location of primary melanoma: Survival differences and sun exposure. J Am Acad Dermatol. 2019;81(2):500–9.

8. Joosse A, Collette S, Suciu S, Nijsten T, Patel PM, Keilholz U, et al. Sex is an independent prognostic indicator for survival and relapse/progression-free survival in metastasized stage III to IV melanoma: a pooled analysis of five European organisation for research and treatment of cancer randomized controlled trials. J Clin Oncol. 2013;31(18):2337–46.

9. Lasithiotakis K, Leiter U, Meier F, Eigentler T, Metzler G, Moehrle M, et al. Age and gender are significant independent predictors of survival in primary cutaneous melanoma. Cancer. 2008;112(8):1795–804.

10. Cancer Genome Atlas N. Genomic Classification of Cutaneous Melanoma. Cell. 2015;161(7):1681–96.

11. InterMEL [Available from: https://intermel.org/about/.

12. Atkins MB, Curiel-Lewandrowski C, Fisher DE, Swetter SM, Tsao H, Aguirre-Ghiso JA, et al. The State of Melanoma: Emergent Challenges and Opportunities. Clin Cancer Res. 2021;27(10):2678–97.

13. Luke JJ, Rutkowski P, Queirolo P, Del Vecchio M, Mackiewicz J, Chiarion-Sileni V, et al. Pembrolizumab versus placebo as adjuvant therapy in completely resected stage IIB or IIC melanoma (KEYNOTE-716): a randomised, double-blind, phase 3 trial. Lancet. 2022.

14. Makunts T, Burkhart K, Abagyan R, Lee P. Retrospective analysis of clinical trial safety data for pembrolizumab reveals the effect of co-occurring infections on immune-related adverse events. PLoS One. 2022;17(2):e0263402.

15. Cheng DT, Mitchell TN, Zehir A, Shah RH, Benayed R, Syed A, et al. Memorial Sloan Kettering-Integrated Mutation Profiling of Actionable Cancer Targets (MSK-IMPACT): A Hybridization Capture-Based Next-Generation Sequencing Clinical Assay for Solid Tumor Molecular Oncology. J Mol Diagn. 2015;17(3):251–64.

16. Zehir A, Benayed R, Shah RH, Syed A, Middha S, Kim HR, et al. Mutational landscape of metastatic cancer revealed from prospective clinical sequencing of 10,000 patients. Nat Med. 2017;23(6):703–13.

17. The R Project for Statistical Computing 2022 [Available from: http://www.R-project.org.

18. Gaffney EF, Riegman PH, Grizzle WE, Watson PH. Factors that drive the increasing use of FFPE tissue in basic and translational cancer research. Biotech Histochem. 2018;93(5):373–86.

19. Vicente A, Bianchini RA, Laus AC, Macedo G, Reis RM, Vazquez VL. Comparison of protocols for removal of melanin from genomic DNA to optimize PCR amplification of DNA purified from highly pigmented lesions. Histol Histopathol. 2019;34(9):1089–96.

20. Satyamoorthy K, Li G, Van Belle PA, Elder DE, Herlyn M. A versatile method for the removal of melanin from ribonucleic acids in melanocytic cells. Melanoma Res. 2002;12(5):449–52.

21. Pujani M, Jain H, Chauhan V, Agarwal C, Singh K, Singh M. Evaluation of Tumor infiltrating lymphocytes in breast carcinoma and their correlation with molecular subtypes, tumor grade and stage. Breast Dis. 2020;39(2):61–9.

22. Millan-Esteban D, Reyes-Garcia D, Garcia-Casado Z, Banuls J, Lopez-Guerrero JA, Requena C, et al. Suitability of melanoma FFPE samples for NGS libraries: time and quality thresholds for downstream molecular tests. Biotechniques. 2018;65(2):79–85.

